# Iliac Vein Compression: A Protective Mechanism Against Symptomatic Pulmonary Embolism Following Lower Extremities Deep Vein Thrombosis

**DOI:** 10.1101/2025.04.12.25325428

**Authors:** Sicheng Yao, Mahebali Jiaerheng, Jiawei Guo, Hongbo Ci

**Affiliations:** Division of Vascular Surgery, People’s Hospital of Xinjiang Uygur Autonomous Region, Urumqi, 830001, People’s Republic of China; Division of Radiology, People’s Hospital of Xinjiang Uygur Autonomous Region, Urumqi, 830001, People’s Republic of China

**Keywords:** Deep vein thrombosis, Iliac vein compression syndrome, Pulmonary embolism, Risk factors

## Abstract

**Objective:** To investigate the protective effect of iliac vein compression in patients with symptomatic pulmonary embolism (PE) of lower extremity deep vein thrombosis (LEDVT).

**Methods:** The clinical data of 82 patients with unilateral LEDVT including computed tomography of pulmonary angiogram (CTPA) and CT image data of common iliac vein from February 2020 to April 2023 were retrospectively analyzed. The patients were divided into two groups according to the presence or absence of PE. Univariate and multivariate logistic regression were used to analyze the risk factors of symptomatic PE in patients with LEDVT. The predictive value of severity of iliac vein compression for symptomatic PE with different measurements were further compared with receiver operating characteristics curve.

**Results:** Univariate analysis showed that compression > 50% (OR = 0.19,95% CI = 0.07-0.51, P = .001) was a protective factor for symptomatic PE. Multivariate analysis showed that compression > 50% (OR = 0.21,95% CI = 0.07-0.64, P = .006) was a protective factor for symptomatic PE. Different compression measurements were compared. With affected caudal diameter as denominator, the area under curve (AUC) was 0.729, the cut-off value was 0.52, the sensitivity was 70.6%, and the specificity was 71.0%. Patent contralateral diameter as denominator, the AUC was 0.729, the cut-off value was 0.49, the sensitivity was 54.9%, and the specificity was 87.1%.

**Conclusions:** In patients with LEDVT, iliac vein compression greater than 50% is a protective factor for symptomatic PE. A larger sample is needed to determine the protective effect of iliac vein compression.

## Introduction

Venous thromboembolism (VTE) includes pulmonary embolism (PE) and deep vein thrombosis (DVT), two pathogenesis represent different clinical manifestations of VTE, constitute a major global burden of cardiovascular disease. Over time, the proportion of PE in VTE has increased while the portion of DVT has decreased[1]. Different and more advanced screening protocols may cause this scenario. PE is reported to occur up to 32% of patients with the lower extremities deep vein thrombosis (LEDVT)[2,3]. The presentation of PE is variable and nonspecific so diagnosis is challenging, most patients with PE are asymptomatic, with only 5% presenting with symptomatic PE[1]. Asymptomatic PE can be noticed until admitted in hospital with long-term sequelae such as chronic thromboembolic pulmonary hypertension (CTEPH)[4,5]. Hence, it is important to identify PE in DVT patients and start early treatments. Computed tomography of pulmonary angiogram (CTPA) is the most common method in PE diagnosis[6]. However, CTPA is not recommended routinely in patients with DVT according to the European Society for Vascular Surgery (ESVS) guidelines due to unclear benefits[6]. Therefore, risk stratification of DVT patients who might progress to PE with the need of CTPA is of paramount importance[7].

In addition to common risk factors that may provoke DVT, it is more likely to occur on the left side, while PE is more likely secondary to right side DVT[8,9]. This may be associated with iliac vein compression, which is more prevalent on the left side due to inherited anatomical positioning[10,11]. Prior to the implementation of IVC filters in clinical practice, IVC ligation was used as a method to prevent PE occurrence. The left side DVT may have been because iliac vein compression acts as a self-filter, preventing the migration of lower extremity thrombi and thereby reducing the likelihood of PE. Previous studies have found that left iliac vein compression greater than 70% increases the risk of DVT, but compression greater than 42.9% or vein diameter less than 6.77 mm decreases the risk of PE[8,12]. However, previous studies have demonstrated inconsistent compression levels and a lack of comparison between different measurements of iliac vein compression[7].

In this study, the relationship between the severity of iliac vein compression and the occurrence of symptomatic PE was further verified by comparing different measurements.

## Materials and Methods

### Study design

A retrospective case-control study was conducted in accordance with the ethical standards of the institutional ethics committee, which waived the requirement for formal approval due to the anonymized and non-interventional nature of the data. All patients provide consent for clinical data research during hospital admission, and further informed consent is waived. At our center, electronic records and image data from the admission date of January 2020 are accessible through a cloud-based data system. Two vascular surgeons independently review medical records to select research participants.

Between February 2020 and April 2023, a total of 1882 patients with a primary diagnosis of DVT were enrolled in this study. Inclusion criteria: patients with LEDVT confirmed by ultrasound or venography who underwent both CTPA and pelvic/iliac vein imaging with or without contrast materials. Exclusion criteria: Unable to determine VTE history association with PE, absence or inability to analyze images, inferior vena cava filter placement prior to PE diagnosis; bilateral DVT. After applying these inclusion criteria, 82 patients were included in the final analysis.

The patients were divided into two groups: those with (n=51) and those without (n=31) PE. In our division, CTPA is only performed on patients with PE-related symptoms. Therefore, all PE cases in this study were defined as symptomatic PE. Symptomatic PE cases had confirmation of a filling defect in the pulmonary arteries on CT angiography with any of the following concurrent clinical symptoms: pleuritic chest pain, shortness of breath, tachypnea, hemoptysis, tachycardia, and hypoxia. The patients’ medical records were reviewed, and demographic information, including age, sex, height, weight, duration of symptoms, in-hospital time, affected limb, and risk factors, was collected. Imaging data on the extent of thrombus, the diameter of the inferior vena cava (IVC), the diameter of the bilateral common iliac veins (CIV), and the status of PE were also obtained.

### Compression percentage measurement

Two board-certified radiologists independently measured the diameter of the IVC and bilateral CIV without being informed of any potential correlation between CIV compression and PE. To evaluate the reliability of the image measurement technique, they used a standardized protocol. For the left side as an example, the minimum CIV diameter was defined as the point of maximal compression where the left common iliac artery crossed the left CIV, while the maximum CIV diameter was defined as 4-6 slices (5mm with each CT slice) distal to that point where there was clear vision of the vessel. The radiologists were masked to each other’s measurements, and the mean values for each vascular diameter obtained in our study represent the average of their reports. To assess the reliability of the measurements, we used intraclass correlation coefficients (ICC). The CIV compression was objectively defined using both qualitative and quantitative measures. These measures were then compared across three distinct compression definitions. A thorough literature review preceded the selection of these three definitions, ensuring that our study was aligned with existing research. The comparison among the three different compression definitions is summarized in Figure 2.

**Figure 1.**
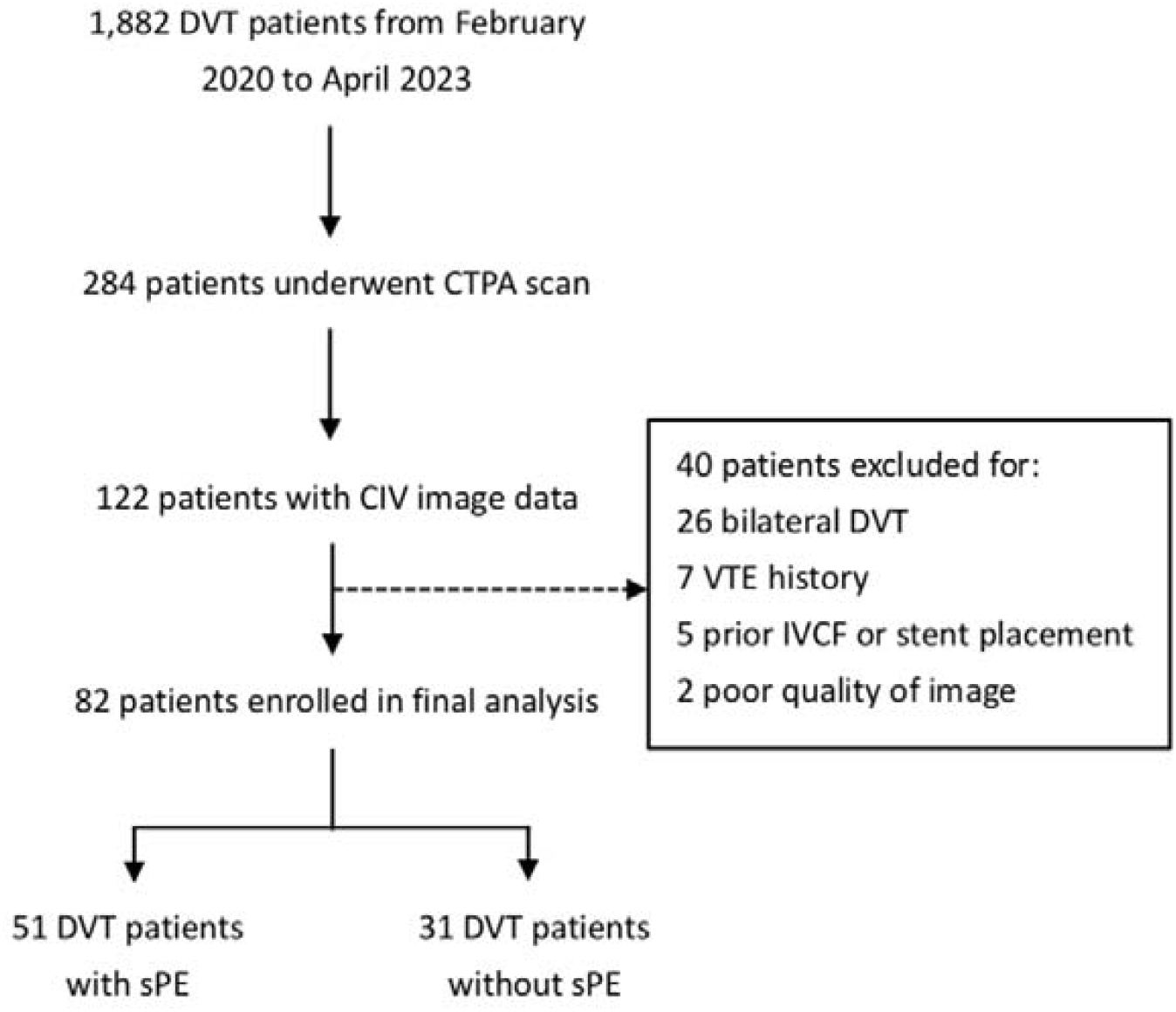
Patients selection flowchart. Abbreviations: DVT, deep vein thrombosis; CTPA, computed tomography of pulmonary angiogram; CIV, common iliac vein; VTE, venous thromboembolism; IVCF, inferior vena cava filter.

**Figure 2.**
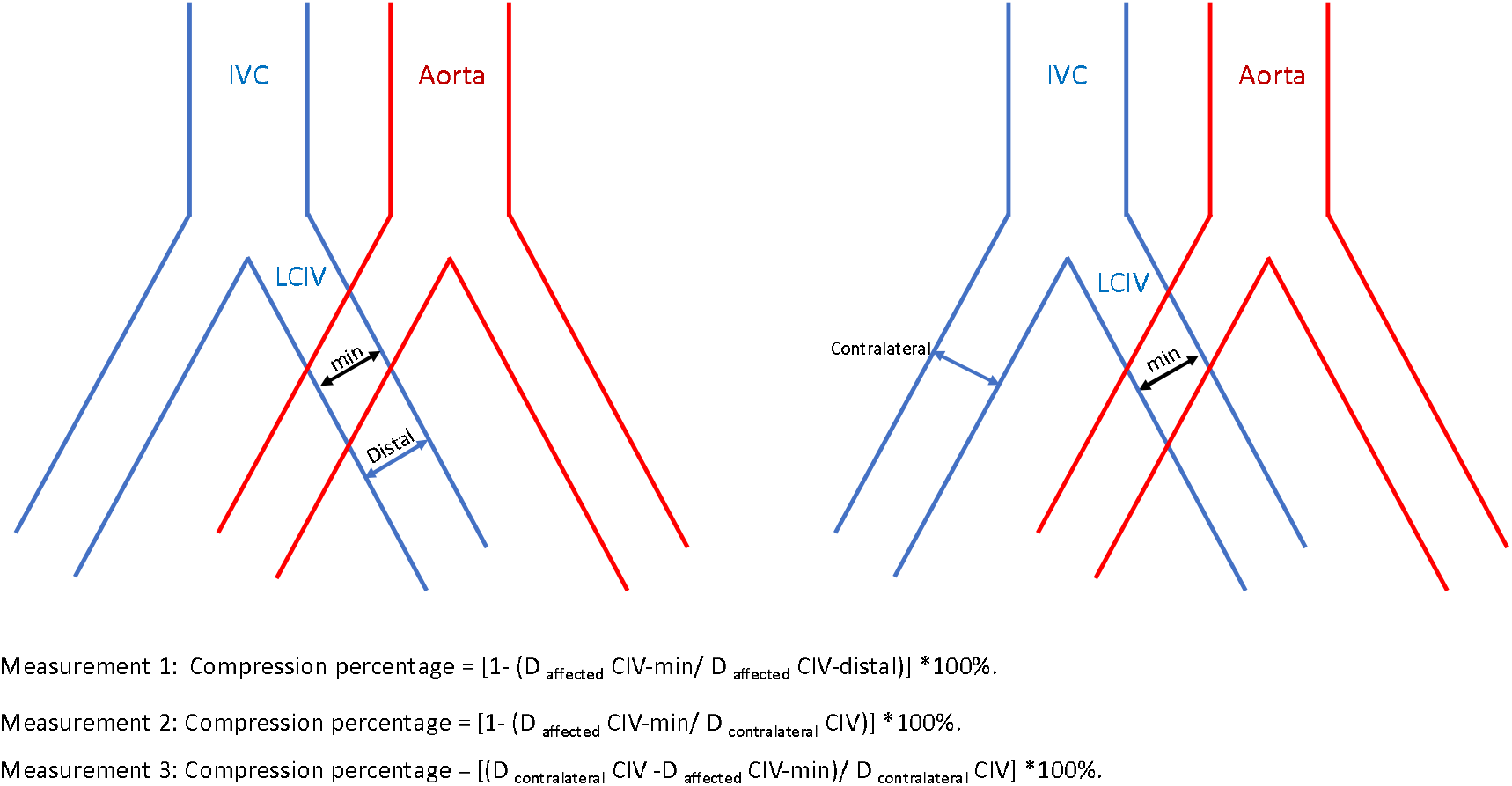
Comparison between different measurements. Abbreviations: IVC, inferior vena cava; LCIV, left common iliac vein; min, minimal diameter.

### Statistical analysis

Baseline demographic variables were described by using means for continuous variables and proportions for categoric variables. For asymmetric data, the median and interquartile range (IQR) were presented. To compare continuous data, the Student t test or Mann-Whitney U test was used. For categorical data, the chi square test or Fisher’s exact test was utilized. We estimated the relationship between CIV compression and symptomatic PE using univariate logistic regression models, providing odds ratios (OR) and 95% confidence intervals (CI). The models were then adjusted for confounders by multivariate logistic regression. The predictive value of CIV compression with different measurements for symptomatic PE was evaluated using receiver operating characteristics (ROC) curves. All statistical analysis was conducted using R software (http://www.R-project.org; R Foundation for Statistical Computing, Vienna, Austria). A *p* value < 0.05 was considered statistically significant.

## Results

### Baseline data

Among 82 patients, demographic data, risk factors and comorbidity between 31 non-symptomatic and 51 symptomatic PE cases were compared. Control and case group share similar mean age (66.00 y vs 62.00 y) and female proportion (64.52% vs 49.02%). Malignancy is the most common risk factors (14.63%). Hypertension, coronary heart disease and diabetes are three most common comorbidities, affecting 30.49%, 14.63%, 12.20% respectively. Patients with symptomatic PE was more likely have a higher weight and BMI (*p* <0.01). Right side DVT patients is more likely to have symptomatic PE compare with left side DVT (*p*<0.01). There were no significant differences in onset time and in-hospital time between two groups. (Table 1)

**Table 1.** Baseline clinical characteristics.

|  | No-SPE (Controls) | SPE (Cases) | Total | *p*-value |
| --- | --- | --- | --- | --- |
|  | (N=31) | (N=51) | (N=82) |  |
| **Demographic data** |  |  |  |  |
| Age, years | 66.00 [48.50;71.00] | 62.00 [54.00;74.00] | 64.00 [49.00;73.00] | 0.924 |
| Female, sex | 20 (64.52%) | 25 (49.02%) | 45 (54.88%) | 0.252 |
| Height, cm | 164.06 ± 8.93 | 167.33 ± 8.33 | 166.10 ± 8.65 | 0.097 |
| Weight, kg | 67.48 ± 12.81 | 76.92 ± 13.52 | 73.35 ± 13.96 | 0.002 |
| BMI, kg/m<sup>2</sup> | 24.97 ± 3.41 | 27.44 ± 4.30 | 26.51 ± 4.15 | 0.008 |
| **Risk factors for venous thromboembolism** |  |  |  |  |
| Recent surgery | 1 (3.23%) | 2 (3.92%) | 3 (3.66%) | 1 |
| Trauma | 2 (6.45%) | 0 (0) | 2 (2.44%) | 0.14 |
| Recent fracture | 3 (9.68%) | 1 (1.96%) | 4 (4.88%) | 0.149 |
| Immobility | 2 (6.45%) | 1 (1.96%) | 3 (3.66%) | 0.553 |
| Malignancy | 5 (16.13%) | 7 (13.73%) | 12 (14.63%) | 0.758 |
| Immune diseases | 0 (0) | 3 (5.88%) | 3 (3.66%) | 0.286 |
| **Comorbidity** |  |  |  |  |
| Heart failure | 2 (6.45%) | 5 (9.80%) | 7 (8.54%) | 0.705 |
| COPD | 1 (3.23%) | 3 (5.88%) | 4 (4.88%) | 1 |
| Coronary heart disease | 2 (6.45%) | 10 (19.61%) | 12 (14.63%) | 0.121 |
| Renal insufficiency | 1 (3.23%) | 2 (3.92%) | 3 (3.66%) | 1 |
| Hypertension | 7 (22.58%) | 18 (35.29%) | 25 (30.49%) | 0.323 |
| Diabetes | 2 (6.45%) | 8 (15.69%) | 10 (12.20%) | 0.305 |
| Cerebral disease | 1 (3.23%) | 3 (5.88%) | 4 (4.88%) | 1 |
| In-hospital time, days | 8.00 [5.50;13.50] | 6.00 [4.00;10.50] | 7.00 [4.00;11.00] | 0.109 |
| Onset time, days | 5.00 [3.00;10.00] | 4.00 [2.00;8.00] | 4.50 [2.50;10.00] | 0.592 |
| **Thrombus distribution** |  |  |  | 0.004 |
| Left | 26 (83.87%) | 26 (50.98%) | 52 (63.41%) |  |
| Right | 5 (16.13%) | 25 (49.02%) | 30 (36.59%) |  |
| Proximal DVT |  |  |  |  |
| Iliofemoral | 24 (77.42%) | 35 (68.63%) | 59 (71.95%) | 0.454 |
| Femoropopliteal | 3 (9.68%) | 12 (23.53%) | 15 (18.29%) | 0.148 |
| Distal DVT | 4 (12.90%) | 4 (7.84%) | 8 (9.76%) | 0.469 |
| **Vascular diameter** |  |  |  |  |
| IVC | 25.04 ± 2.95 | 26.11 ± 2.57 | 25.71 ± 2.75 | 0.089 |
| LCIV-min | 4.43 [3.50;5.32] | 5.63 [4.53;7.13] | 4.87 [3.97;6.62] | 0.001 |
| Distal LCIV | 9.73 ± 2.78 | 10.73 ± 3.22 | 10.35 ± 3.08 | 0.158 |
| RCIV-min | 6.80 ± 2.12 | 7.44 ± 2.50 | 7.20 ± 2.37 | 0.237 |
| Distal RCIV | 11.10 ± 2.11 | 11.56 ± 2.69 | 11.38 ± 2.48 | 0.418 |

Abbreviations: SPE, symptomatic pulmonary embolism; COPD, chronic obstructive pulmonary diseases; DVT, deep vein thrombosis; IVC, inferior vena cava; LCIV, left common iliac vein; RCIV, right common iliac vein.

### Measurement of iliac vein compression

For the main qualitative measurements showed in Table 2, ICC ranged from 0.715 to 0.859. Mean diameter of IVC, distal LCIV, minimal RCIV and distal RCIV were 25.71 mm, 10.35 mm, 7.20 mm, 11.38 mm respectively. The median minimal diameter of LCIV was 4.87 mm. Patients without symptomatic PE have a smaller minimum LCIV diameter (4.43 mm vs 5.63 mm). (Table 2)

**Table 2.** Results of CT readings.

|  | **Reader 1 (mm)** | **Reader 2 (mm)** | **Mean\* (mm)** | **ICC** |
| --- | --- | --- | --- | --- |
| IVC | 25.33±3.14 | 26.08±2.79 | 25.7 | 0.715 |
| LCIV-min | 5.71±1.92 | 5.05±2.50 | 5.38 | 0.78 |
| Distal LCIV | 10.71±2.85 | 9.99±3.50 | 10.35 | 0.859 |
| RCIV-min | 6.69±2.20 | 7.71±2.82 | 7.2 | 0.76 |
| Distal RCIV | 11.65±2.49 | 11.11±2.76 | 11.38 | 0.786 |

\*Mean diameter represent the average of two radiologists' readings.
Abbreviations: IVC, inferior vena cava; LCIV, left common iliac vein; RCIV, right common iliac vein; ICC, Intraclass Correlation Coefficients.

### Quantitative Analysis

Initially, the univariable analysis indicated that an increased CIV diameter is a significant risk factor for symptomatic PE, with the increased minimum diameter of CIV associated with a 68% increase in the odds of developing symptomatic PE (Odds Ratio [OR] 1.68, 95% Confidence Interval [CI] 1.22-2.33). In light of existing literature, we hypothesized that a CIV diameter larger than 6.4 mm would correlate with greater odds of symptomatic PE.

However, this hypothesis was not statistically supported and prompted a revision of the cut-off point to 6.0 mm. Upon reanalysis, data revealed that a CIV diameter less than 6.0 mm was associated with a significantly reduced incidence of symptomatic PE (OR 0.13, 95% CI 0.04-0.48). After adjusting for sex, BMI, and the limb affected, the protective effect of a smaller CIV diameter (<6.0 mm) remained strong, with an 87% reduction in symptomatic PE odds (OR 0.13, 95% CI 0.03-0.55). (Table 3)

**Table 3.** Quantitative analysis results between pulmonary embolism (PE) and common iliac vein (CIV) compression.

| Analysis | LCIV minimal diameter (mm) |  |  |  |
| --- | --- | --- | --- | --- |
|  | ≤ 6.4 | > 6.4 | ≤ 6.0 | > 6.0 |
| Unadjusted | NA | NA | 0.13 (0.04-0.48, p=.002) | 7.67 (2.06-28.48, p=.002) |
| adjusted* | - | - | 0.13 (0.03-0.55, p=.005) | 7.48 (1.81-30.93, p=.005) |

\*adjusted for sex, BMI, DVT side.
Presented with ORs, with 95% CIs.
NA = not applicable.

### Qualitative analysis

CIV compression degree was further used to analysis the association qualitatively. Different knots under three different measurements were analyzed, four levels with a separate 25% compression was initiated but several groups had insufficient cases for statistical analysis.

Univariate analysis showed that, right lower extremity (OR = 5.00,95% CI = 1.66-5.07, P = .004), BMI (OR = 1.17,95% CI = 1.04-1.32, P = .011), and CIV compression ≤ 50% (OR = 5.29,95% CI = 1.96-14.16, P = .001) were risk factors for symptomatic PE, compression > 50% (OR = 0.19,95% CI = 0.07-0.51, P = .001) was a protective factor for symptomatic PE. Multivariate analysis after adjusted for side, BMI and sex showed that compression ≤50% (OR = 4.67,95% CI = 1.57-14.08, P = .006) was a risk factor for symptomatic PE, and compression > 50% (OR = 0.21,95% CI = 0.07-0.64, P = .006) was a protective factor for symptomatic PE. (Table 4) ROC curves were then applied to analysis consistent between different measurements, one measurement was removed due to unreliable results. Two measure protocols were enrolled in final analysis. With affected caudal diameter as denominator, the AUC was 0.729, the cut-off value was 0.52, the sensitivity was 70.6%, and the specificity was 71.0%. Patent contralateral diameter as denominator, the AUC was 0.729, the cut-off value was 0.49, the sensitivity was 54.9%, and the specificity was 87.1%. (Figure 3)

**Table 4.** Qualitative analysis results between pulmonary embolism (PE) and common iliac vein (CIV) compression.

| Analysis | Measurement 1 |  | Measurement 2 |  | Measurement 3 |  |
| --- | --- | --- | --- | --- | --- | --- |
|  | percentage ≤ 50% | 50% < percentage | percentage ≤ 50% | 50% < percentage | percentage ≤ 50% | 50% < percentage |
| Unadjusted | 5.27 (1.96-14.16, p=.001) | 0.19 (0.07-0.51, p=.001) | 8.22 (2.51-26.91, p<.001) | 0.12 (0.04-0.40, p<.001) | 0.76 (0.28-2.07, p=.592) | 0.37 (0.11-1.30, p=.121) |
| adjusted* | 4.71 (1.57-14.08, p=.006) | 0.21 (0.07-0.64, p=.006) | 6.66 (1.82-24.32, p=.004) | 0.15 (0.04-0.55, p=.004) | NA | 0.58 (0.15-2.25, p=.436) |

\*adjusted for sex, BMI, DVT side.
Presented with ORs, with 95% CIs.
NA = not applicable.
Measurement 1: Compression percentage = $[1 - (D_{\text{affected CIV-min}} / D_{\text{affected CIV-distal}})] * 100\%$ .
Measurement 2: Compression percentage = $[1 - (D_{\text{affected CIV-min}} / D_{\text{contralateral CIV}})] * 100\%$ .
Measurement 3: Compression percentage = $[(D_{\text{contralateral CIV}} - D_{\text{affected CIV-min}}) / D_{\text{contralateral CIV}}] * 100\%$ .

**Figure 3.**
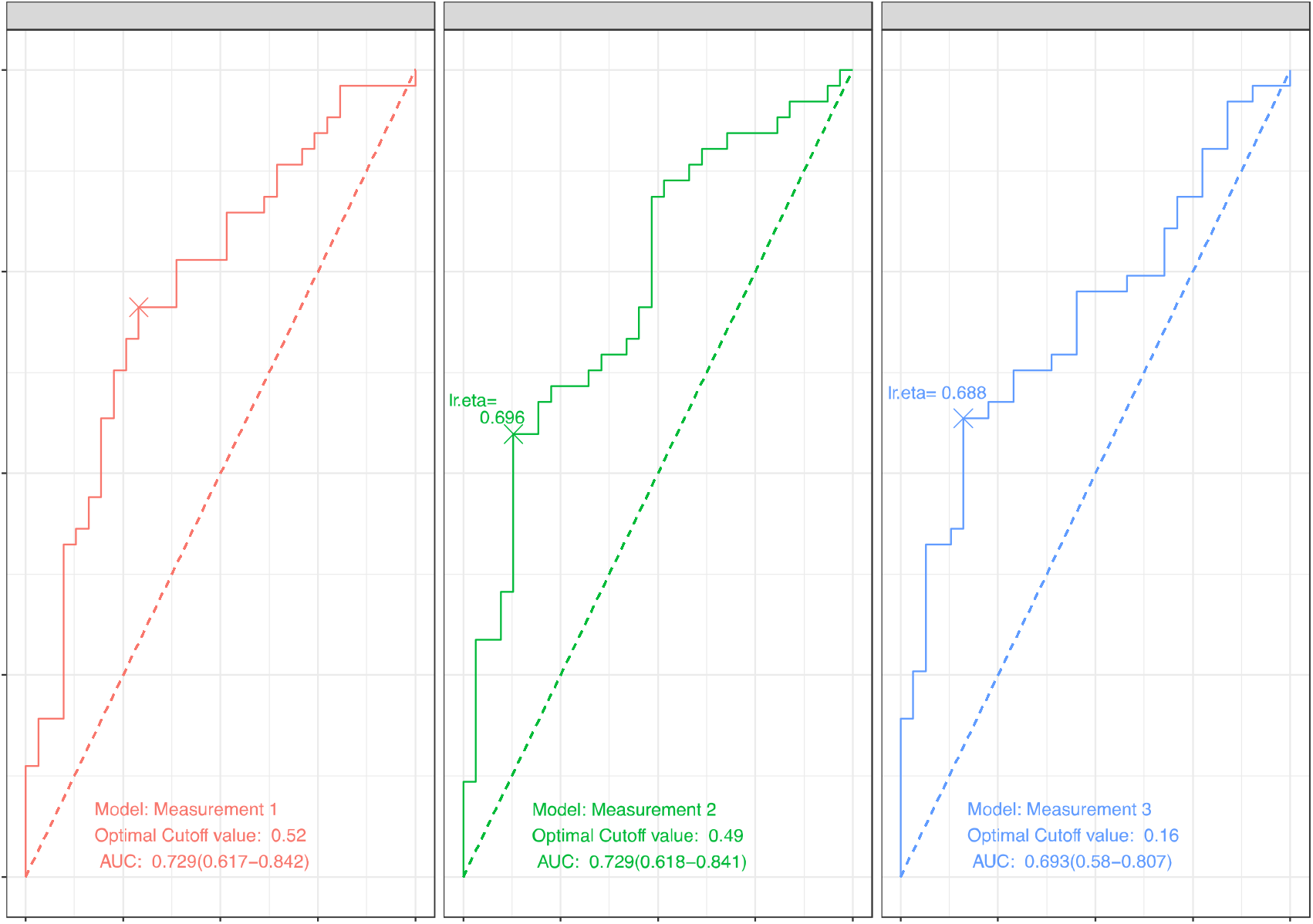
Receiver operating characteristics curves with different measurements. Abbreviations: Sens, sensitivity; Spec, specificity; PPV, positive predictive value; NPV, negative predictive value, AUC, area under curve.

## Discussion

Many risk factors have been identified as important in the development of DVT, but clinical risk stratification related to DVT progressing to PE remains challenging. Our study found that, among patients with DVT, CIV compression greater than 50% serves as a protective factor against symptomatic PE. Two measure protocols used in our study have a similar result and both show a reduce risk of symptomatic PE in patients with CIV compression greater than 50%.

The association between iliac vein compression and DVT have been well elucidated. Both traditional observational study and three-dimensional reconstruction research showed iliac vein compression greater than 70% was associated with substantially greater odds for left DVT[8,13–15]. However, the character of iliac vein compression in the progression of DVT to PE is not fully understand. Silent PE occurs in approximately one third of patients with deep venous thrombosis[3]. And PE develops more frequently in patients with right sided DVT than in those with left sided DVT[16,17]. Similar with previous case-control study, we also found that patients with right DVT (25 of 30) were more likely to have a history of PE than patients with left DVT (26 of 52). Hou et al found patients with typical PE manifestations showed a higher proportion of patients with right lower extremity DVT than left lower extremity DVT (26.7% vs. 17.7%, P = 0.035)[17]. Higher symptomatic PE proportion in our study could related to the cases and controls had been considered high risk of PE and CTPA was used to confirm PE diagnosis.

DVT patients with concomitant iliac vein compression have a decreased risk of PE compared with those without compression[18]. Which means the association between laterality and PE could be explained by the compression caused by May-Turner syndrome. Previous studies have found that CIV compression could act as a physical barrier to limit thrombus migration[10]. Keith et al analyzed 75 unilateral DVT patients and they found that those with an ipsilateral CIV lumen of 4 mm or less have an 83% lower risk of developing symptomatic PE compared with patients with a CIV lumen greater than 4 mm[10]. In their study, minimum CIV diameters were measured 1 cm below the IVC bifurcation on CT images[10]. A recent study by Shi et al found that a CIV diameter of < 6.77 mm or a compression percentage > 42.9% were protective factors against PE[12]. Measurement of CIV compression percentage in their article was evaluated by the diameter at the point of maximum compression divided by the minimum diameter of the CIV caudal to the compression[12]. The contralateral distal CIV diameter was used as the reference denominator for patients with thrombus involving the CIV in their study[12]. This study found compression > 50% (OR = 0.21,95% CI = 0.07-0.64, P = .006) was a protective factor for symptomatic PE after adjusted for side, BMI and sex. Shi et al. categorized compression between 25-50% as mild compression and did not observe a significant reduction in PE risk[12]. Our results are consistent with these findings, and we further observed that compression less than 50% is associated with a higher risk of PE compared to the group with more compression.

This study found that a CIV diameter less than 6.0 mm was associated with a lower incidence of symptomatic PE (adjusted odds ratio [adOR] 0.13, 95% confidence interval [CI] 0.03-0.55). Additionally, the mean diameters of the distal left CIV and right CIV were 10.35mm and 11.38mm, respectively. Assuming an average distal diameter of 11mm and a 50% compression of the vessel, the resulting diameter would be 5.5mm. Due to the small sample size, the results of the quantitative and qualitative analyses were limited but they do show similar results. Different with previous research, this study further investigated the potential influence of different measure protocols. Two measurements in the final analysis both used the minimal diameter at the point of compression but differed in the denominator. We hypothesized that using the affected limb’s caudal diameter as the denominator could underestimate the original luminal diameter of the CIV and consequently overestimate the compression percentage. Using the patent contralateral CIV diameter as an internal control might produce different results. Results showed that with affected caudal diameter as the denominator, an Area Under the Curve (AUC) of 0.729 was achieved, indicating a moderate level of discriminative ability. The respective cut-off value of 0.52 offered 70.6% sensitivity and 71.0% specificity. Using the patent contralateral diameter as the denominator yielded a similar AUC but provided a different trade-off between sensitivity and specificity. Both protocols can be used in the measurement of CIV compression, but further studies with larger sample sizes are required.

Based on the results of our analyses, we did not find any statistically significant differences in the effects of sex or DVT levels (proximal and distal) on PE risk. We also note that distal DVT were typically treated with oral anticoagulation in our division, and no CT scans were performed if patients presented without positive PE symptoms. This may have led to some asymptomatic PE patients being missed. Additionally, ethnicity is another factor that could contribute to different results in previous studies. The studies by Narayan et al and Chan et al were conducted in the US, and the reported distal LCIV diameters (11.4mm and 11.2mm, respectively) were larger than the 10.35mm diameter reported in our study using the same measurement protocol[8,10]. Additional research is needed to further explore these findings.

There are other limitations that must be considered when interpreting its findings. Firstly, the retrospective nature of the study introduces an unavoidable selection bias, as only patients with available iliac vein CT images and CTPA data were included, while individuals with a bilateral or prior history of VTE were excluded. Additionally, CTPA is not routinely performed in our division, but rather reserved for patients at high risk of PE or with strong suspicions of PE. This practice may introduce a bias towards the identification of more severe cases. Furthermore, some CIV images were obtained through pelvic or abdominal scans, either with or without contrast materials. Direct injection of contrast materials can dilate the iliac vein, potentially biasing the true luminal diameter measurement. To ensure accurate measurements, future studies should use consistent image acquisition techniques. An immortal time bias also exists in this study, as no follow up was conducted. This means that some DVT patients may have developed PE after discharge from hospital, but it is difficult to document the full natural history of VTE in these cases. Lastly, diagnosed DVT patients receive anticoagulation therapy immediately following a bleeding risk evaluation. This therapy can prevent the progression of small pulmonary emboli, potentially reducing their incidence.

In conclusion, iliac vein compression, particularly at levels greater than 50%, appears to reduce the risk of PE in patients with DVT. The consistency of findings across two measurement protocols further supports the reliability of these results. Further studies are needed for larger sample size, standard measurements and long term of follow up. The protective effect of iliac vein compression may influence future clinical risk stratification and management strategies in DVT patients.

## Data Availability

All data produced in the present study are available upon reasonable request to the authors.

## Author contribution

S.C. Yao was contributed to the design and implementation of the statistical work, and drafted the manuscript. H.B. Ci was a contributor to the clinical work and revised the manuscript. M. Jiaerheng and J.W. Guo were radiologists contributed to image analysis.

